# The Essence of Nursing Competence: A Philosophical Inquiry into Clinical Performance and Professional Autonomy

**DOI:** 10.1101/2025.09.07.25335259

**Authors:** Arief Yanto, Moses Glorino Rumambo Pandin, Menik Kustriyani

## Abstract

**Background:** Nursing competence is a multifaceted construct that extends beyond technical skill to encompass situated knowing, ethical discernment, and intentional action. In contemporary healthcare, the interplay between clinical performance and professional autonomy shapes the quality of patient-centred care.

**Purpose:** This article philosophically investigates the essence of nursing competence, aiming to elucidate how autonomy functions as a foundational pillar for competent, ethically grounded practice.

**Methods:** A philosophical inquiry employing interpretive phenomenology and critical reflective practice was undertaken. Clinical narratives and professional experiences were systematically analysed to uncover the relational and contextual dimensions of competence.

**Results:** Findings reveal that authentic nursing competence is dynamic rather than static, emerging from a continuous dialectic between expert practice and moral agency. Autonomy enables nurses to transcend checklist-driven tasks, fostering presence, relational depth, and contextual judgment. The study highlights the risk of reduced autonomy leading to technocratic care, thereby compromising holistic, patient-centered outcomes.

**Conclusion:** Professional autonomy is indispensable for the development and enactment of true nursing competence. Educational models and health policies should therefore prioritize critical thinking, moral courage, and intellectual independence to nurture competent practitioners capable of navigating complex clinical landscapes.

## Introduction

A philosophy-based approach to evaluating nursing performance is essential because it facilitates a deeper understanding and enhancement of nursing practice, encompassing not only technical aspects but also humanistic, ethical, and reflective dimensions. Philosophy emphasizes that nurses should regard patients as whole persons rather than merely medical care objects, attending to physical, psychological, social, and spiritual aspects. This approach reinforces humanitarian values, empathy, and caring within nursing practice (Agustini et al., 2023; Cheraghi et al., 2019; Tembo, 2024).

Nurses’ reflection on clinical experiences can cultivate practical wisdom (phronesis) and enhance the quality of clinical decision-making (Corcoran and Cook, 2023; Rentmeester and Liebzeit, 2023). Understanding and internalizing core nursing values such as caring, patient autonomy, and justice can shape professional identity and boost nurses’ motivation and self-confidence (Suandika et al., 2021). Complex challenges, uncertainties, and changes in health care demand critical and creative thinking to keep nursing performance relevant and adaptive (Bender et al., 2021; Dolan et al., 2022; Krol et al., 2024).

This study aims to conduct a philosophical reflection on the essence of nursing competence, focusing specifically on two central aspects: clinical performance and professional autonomy. Clinical performance is not merely the ability to execute tasks; it is an expression of phronesis practical wisdom cultivated through experience, empathy, and ongoing reflection on patient context. Meanwhile, professional autonomy is not just a right or authority but a manifestation of ethical responsibility, mastery of knowledge, and the capacity to make accountable decisions in complex and uncertain situations.

The article presents a critical review integrating philosophical perspectives from various nursing traditions, including phenomenology, caring theory, and feminist epistemology. Through a synthesis of recent scholarly literature (2021–2025), the article explores how nursing competence is not merely something performed, but something constituted through relationships, reflection, and meaningful moral presence.

The purpose of this manuscript is to reveal the philosophical dimensions of nursing competence that are often overlooked in practice measurement standards; to analyse the role of professional autonomy as a pillar in fostering ethical and sustainable clinical performance; and to propose the integration of nursing philosophy into education, policy, and everyday practice to reinforce a cohesive professional identity. This article is expected to constitute an intellectual contribution to the development of nursing as a discipline that is not only scientific but also humane, critical, and rooted in profound values.

## Method

This research employed a literature-review approach. The databases searched were Scopus, Web of Science, PubMed, ScienceDirect, and Dimensions. Literature was retrieved using the following keyword string: *“nursing competence OR clinical performance OR professional autonomy AND philosophy of nursing OR philosophical inquiry OR phenomenology OR caring theory OR feminist epistemology OR phronesis OR practical wisdom OR embodied knowledge OR ethical decision-making.”*

Inclusion criteria were: articles published in English between 2021 and 2025, full-text availability, and relevance to the themes of nursing competence, philosophical inquiry into clinical performance, and professional autonomy. Exclusion criteria were: publications older than five years, review articles, and studies published in Indonesian.There were 82 articles from the Scopus database, 21 articles from Web of Science database, 227 articles from Sciencedirect database, 520 articles from PubMed database, and 28 articles from Dimensions, so the total articles obtained are 878 articles. After screening based on inclusion and exclusion criteria, 15 research articles were obtained consisting international articles.

**Table 1.**
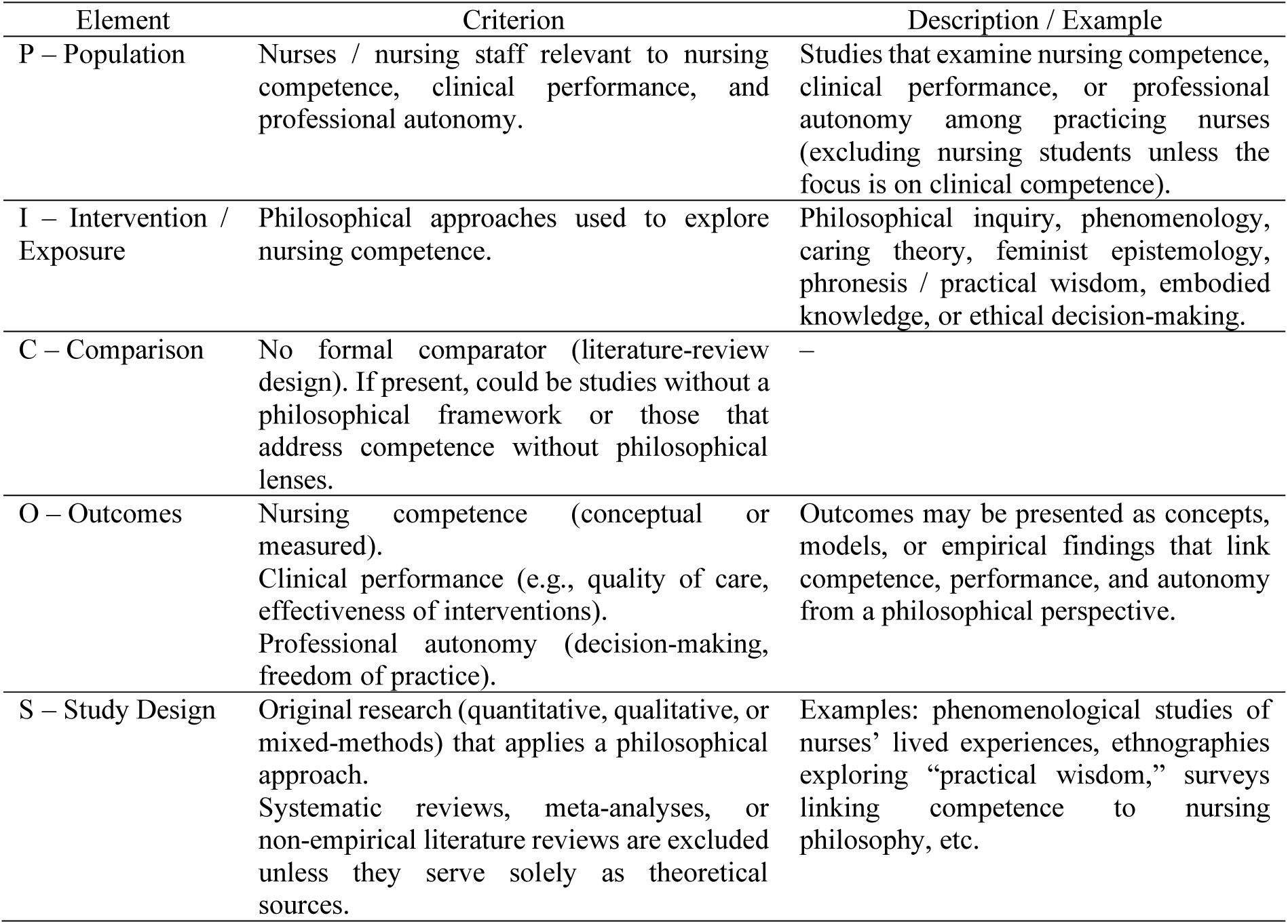
PICOS Table for Article Screening.

## Results

This section presents a comprehensive synthesis of findings derived from the philosophical analysis of nursing competence, with a particular focus on its interdependence with clinical performance and professional autonomy. The synthesis is structured around key thematic dimensions extracted from the narrative accounts and reflective insights of practicing nurses. These dimensions ranging from embodied knowing and ethical judgment to relational presence and institutional constraints reflect the complex, dynamic nature of competent nursing practice. By organizing the data into a structured framework, the synthesis table aims to clarify how autonomy is not merely a policy concern but an ontological condition for authentic nursing.

**Table 1.**
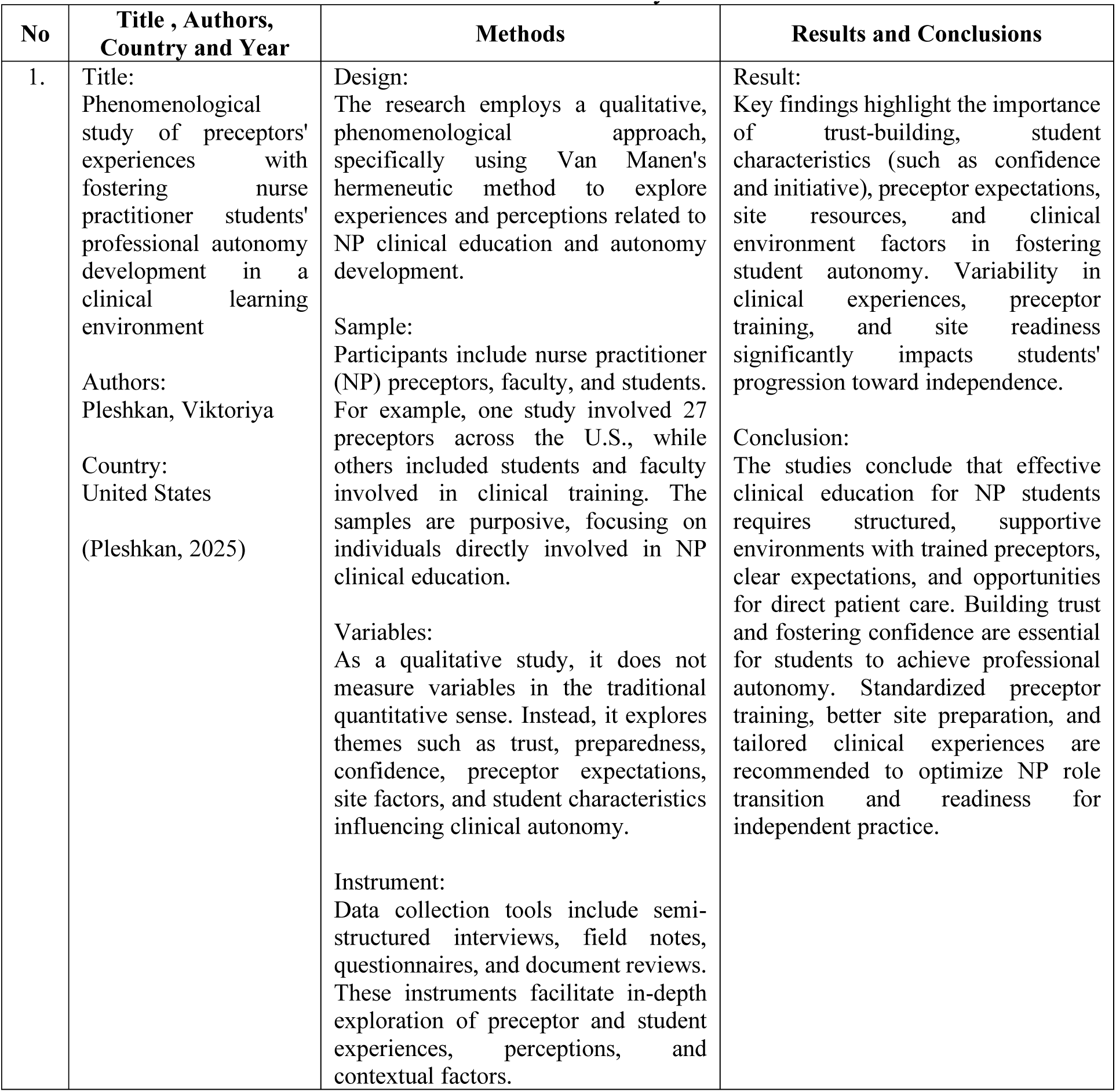
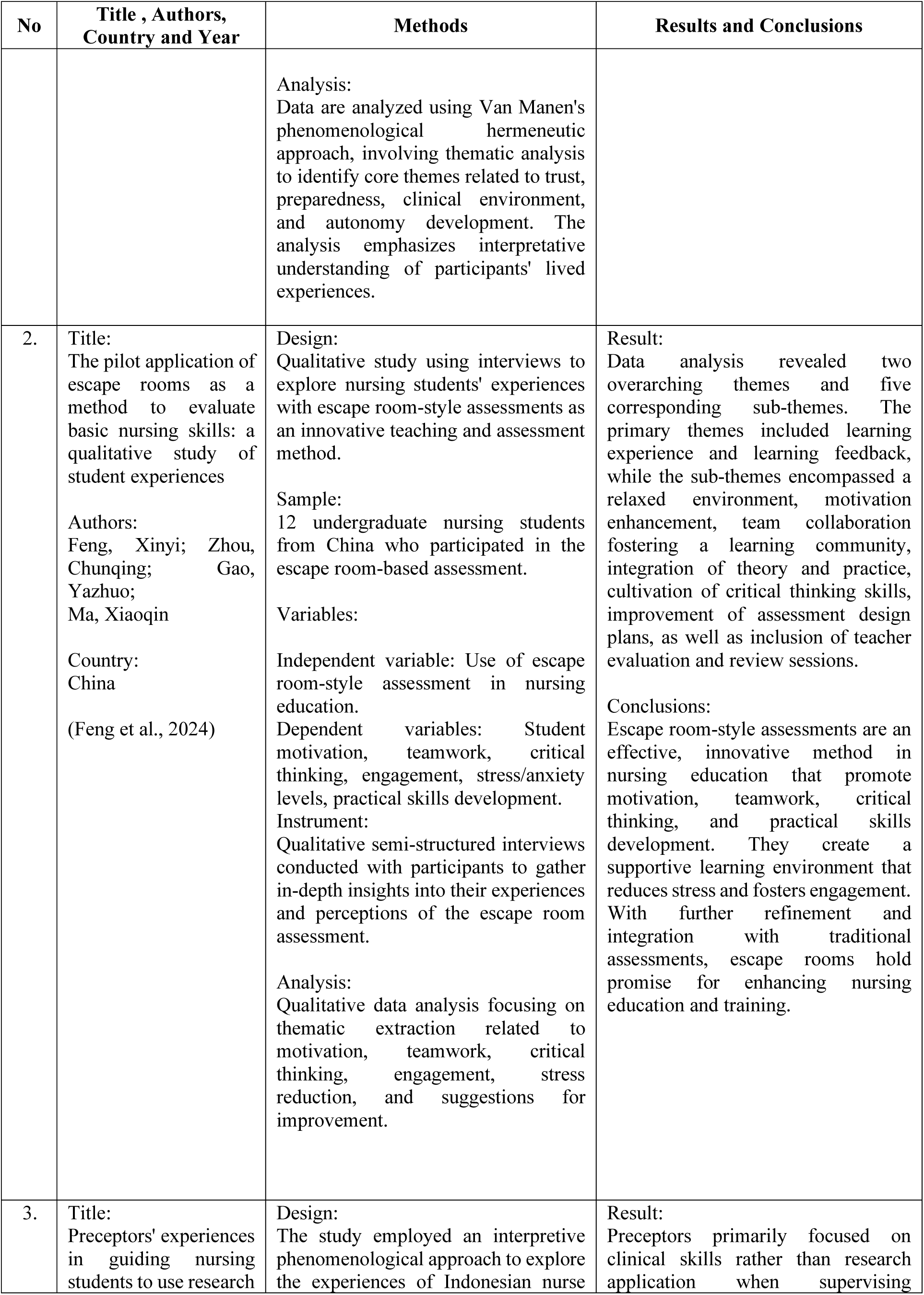
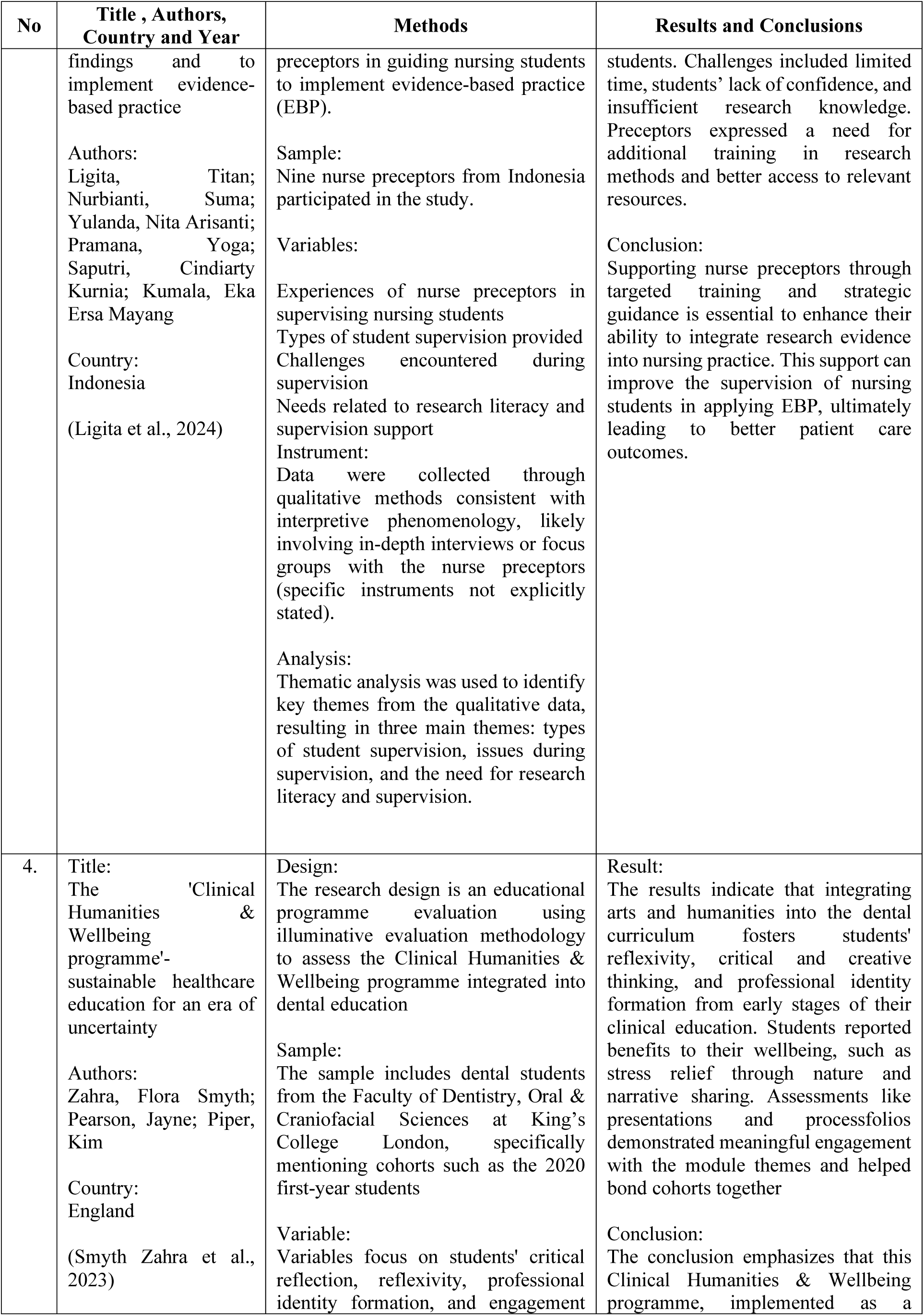
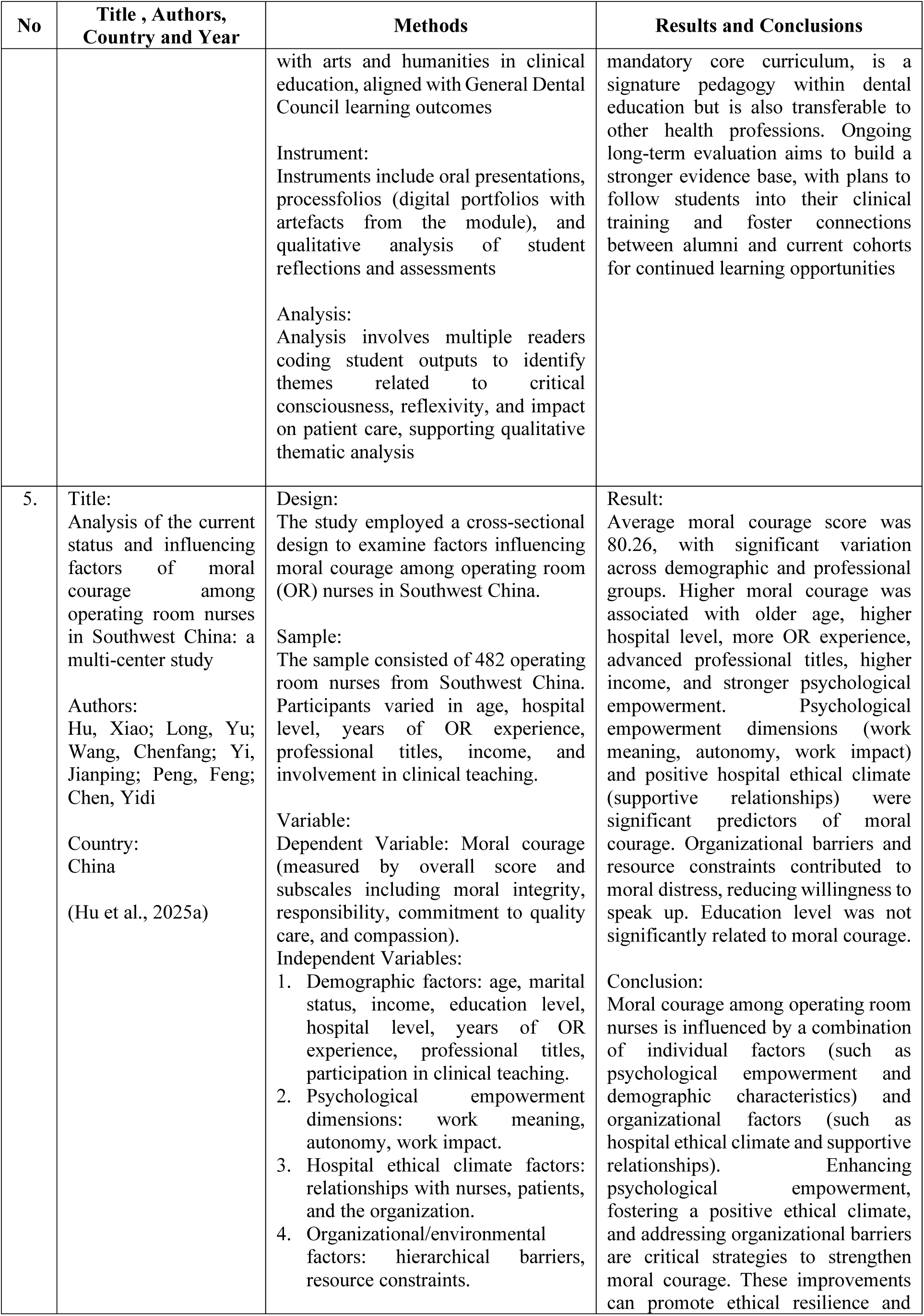
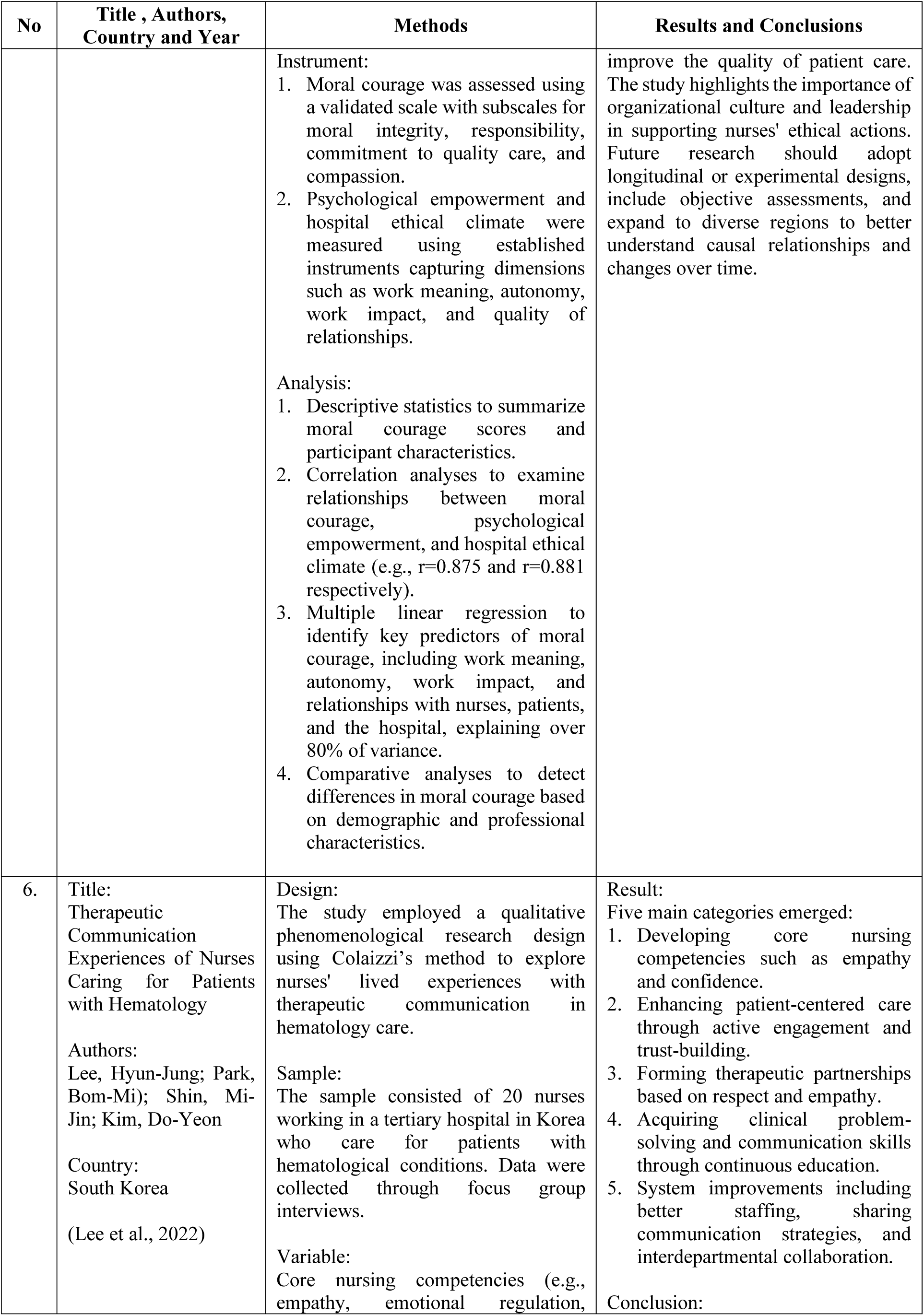
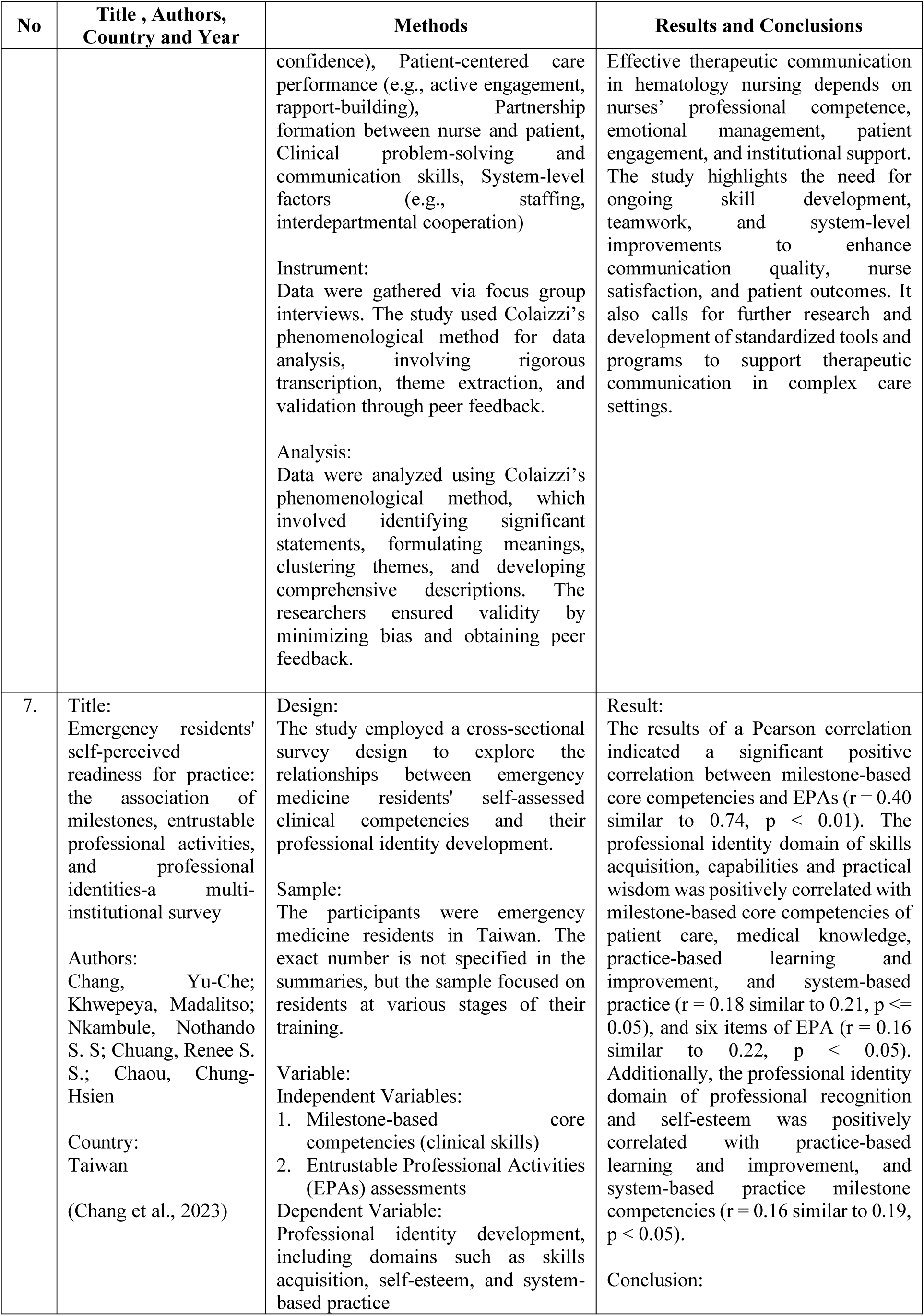
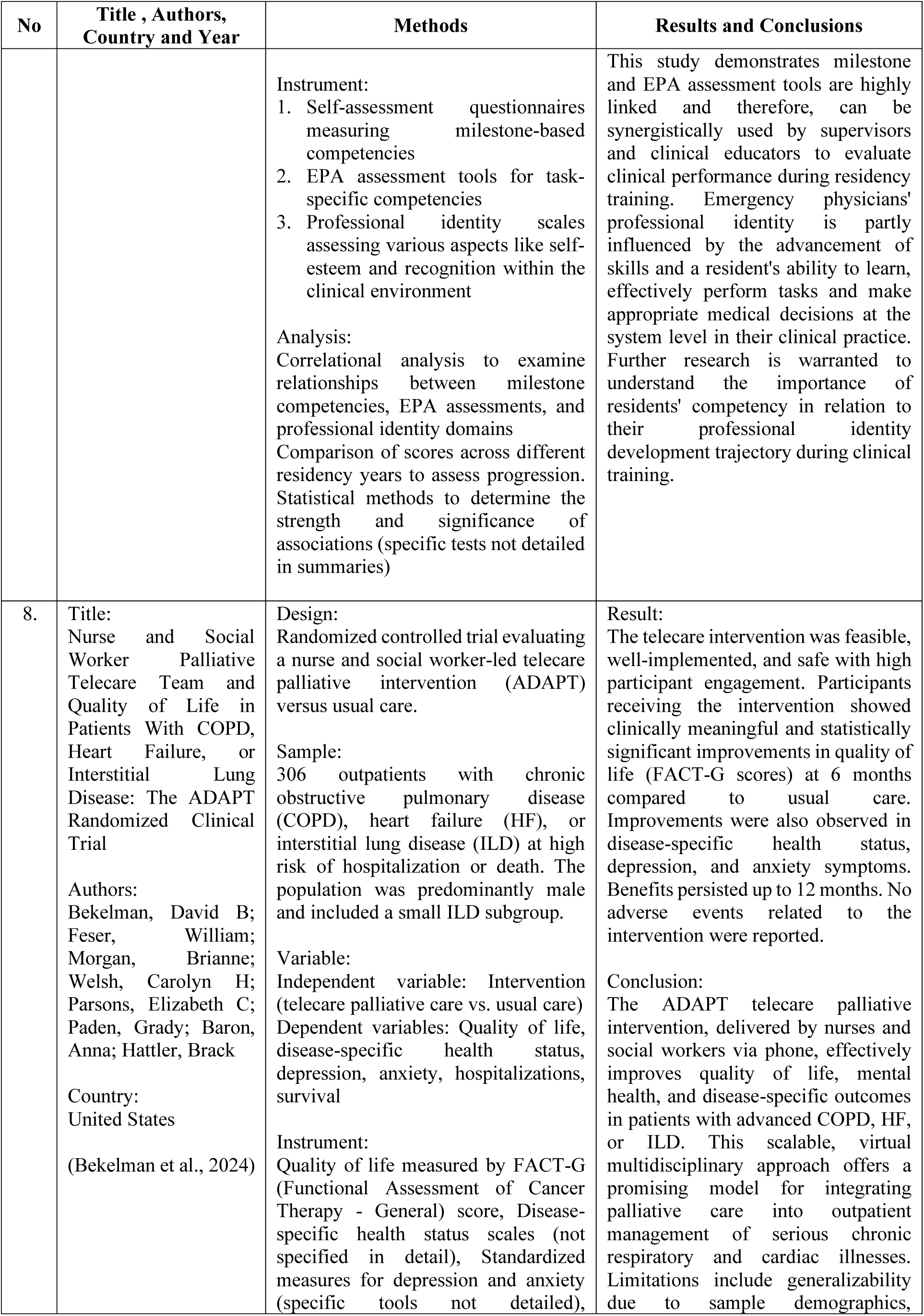
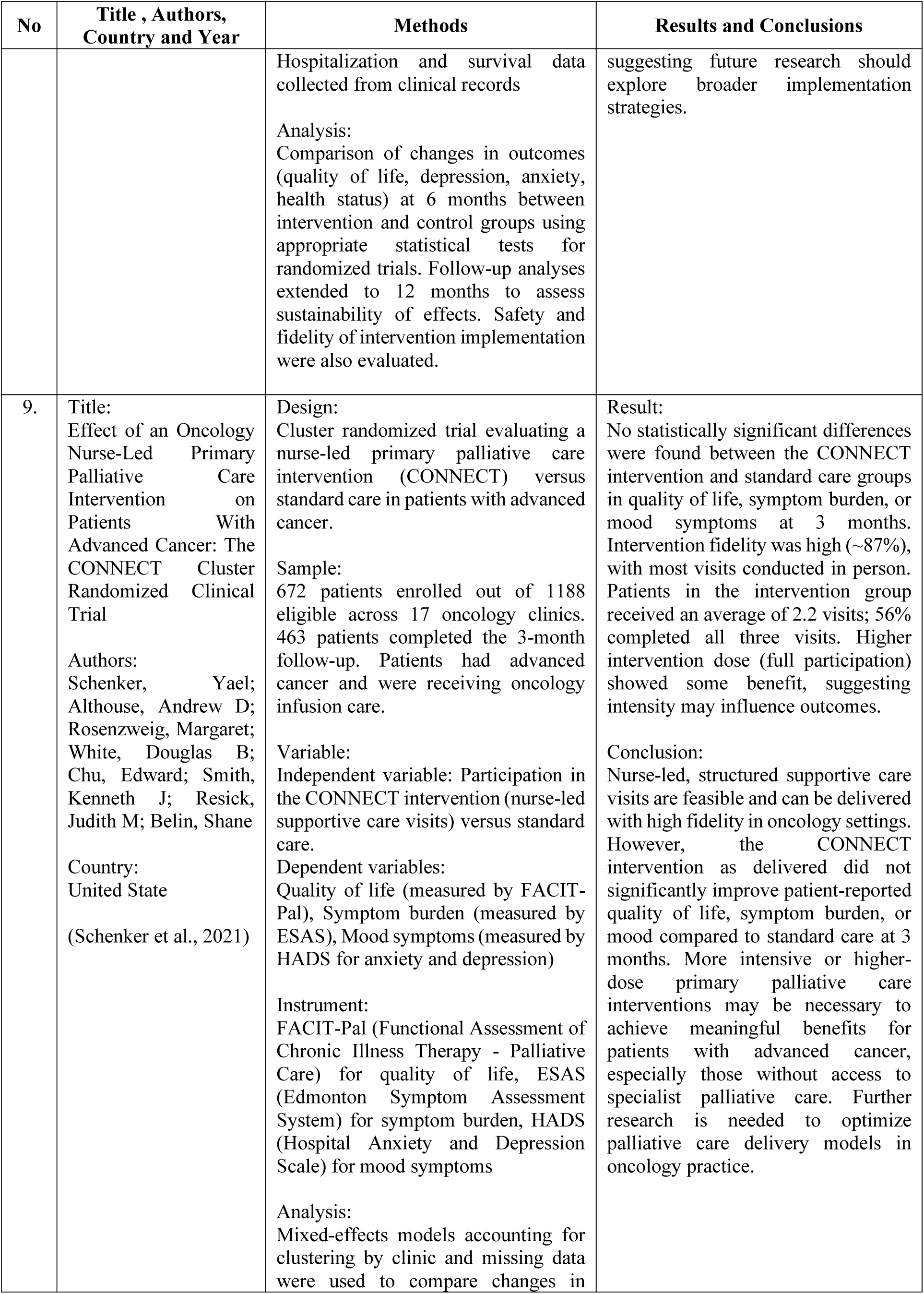
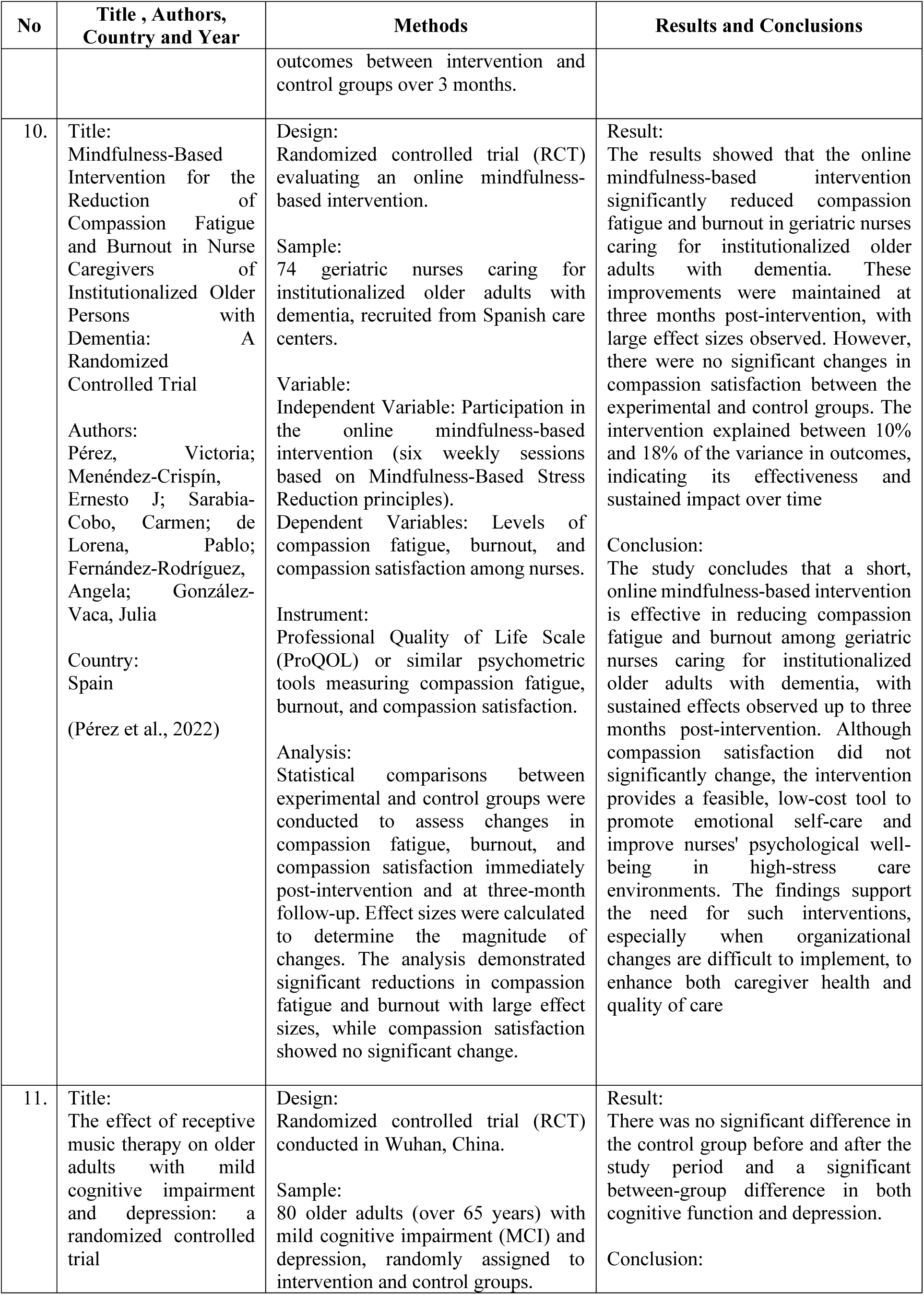
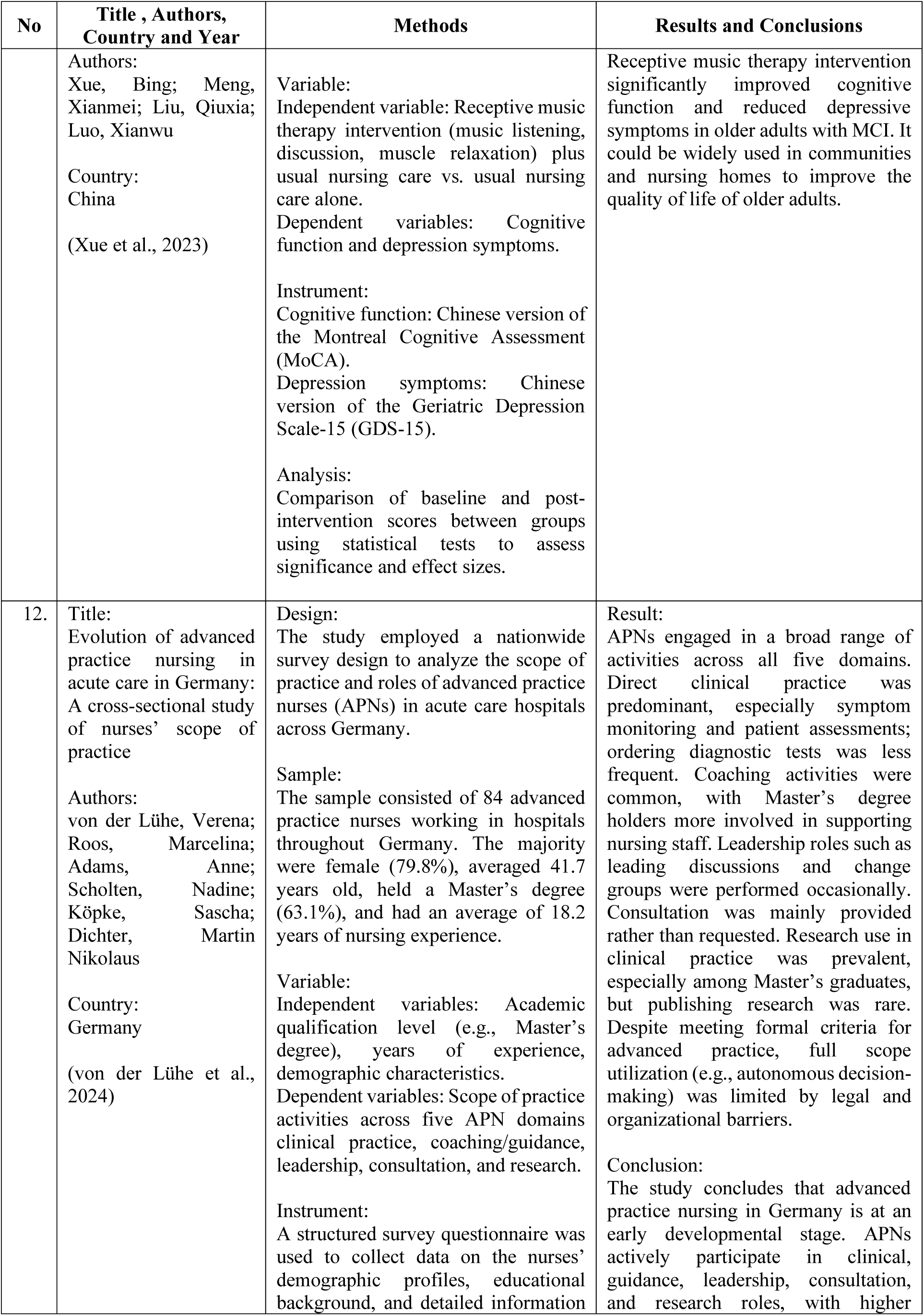
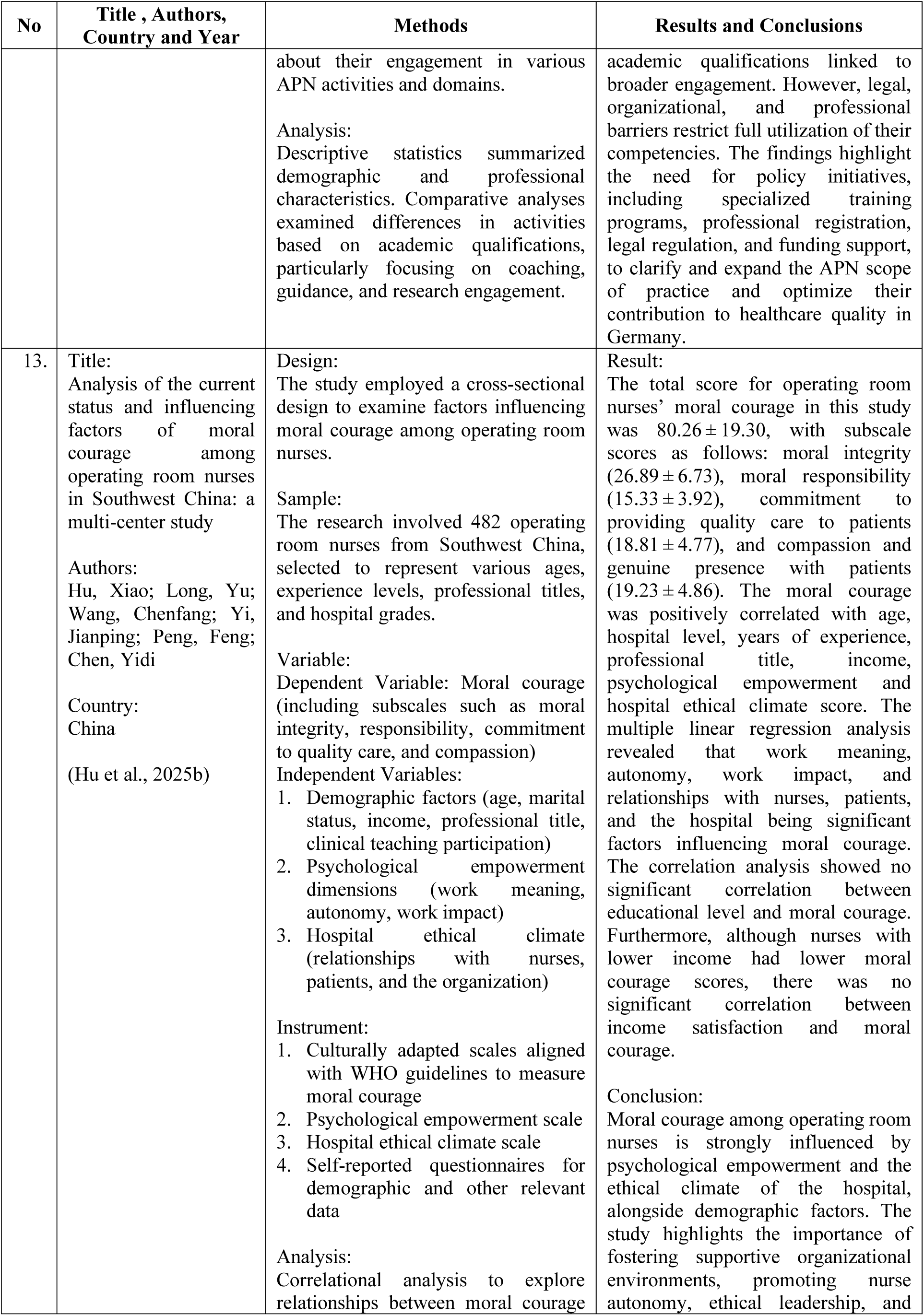
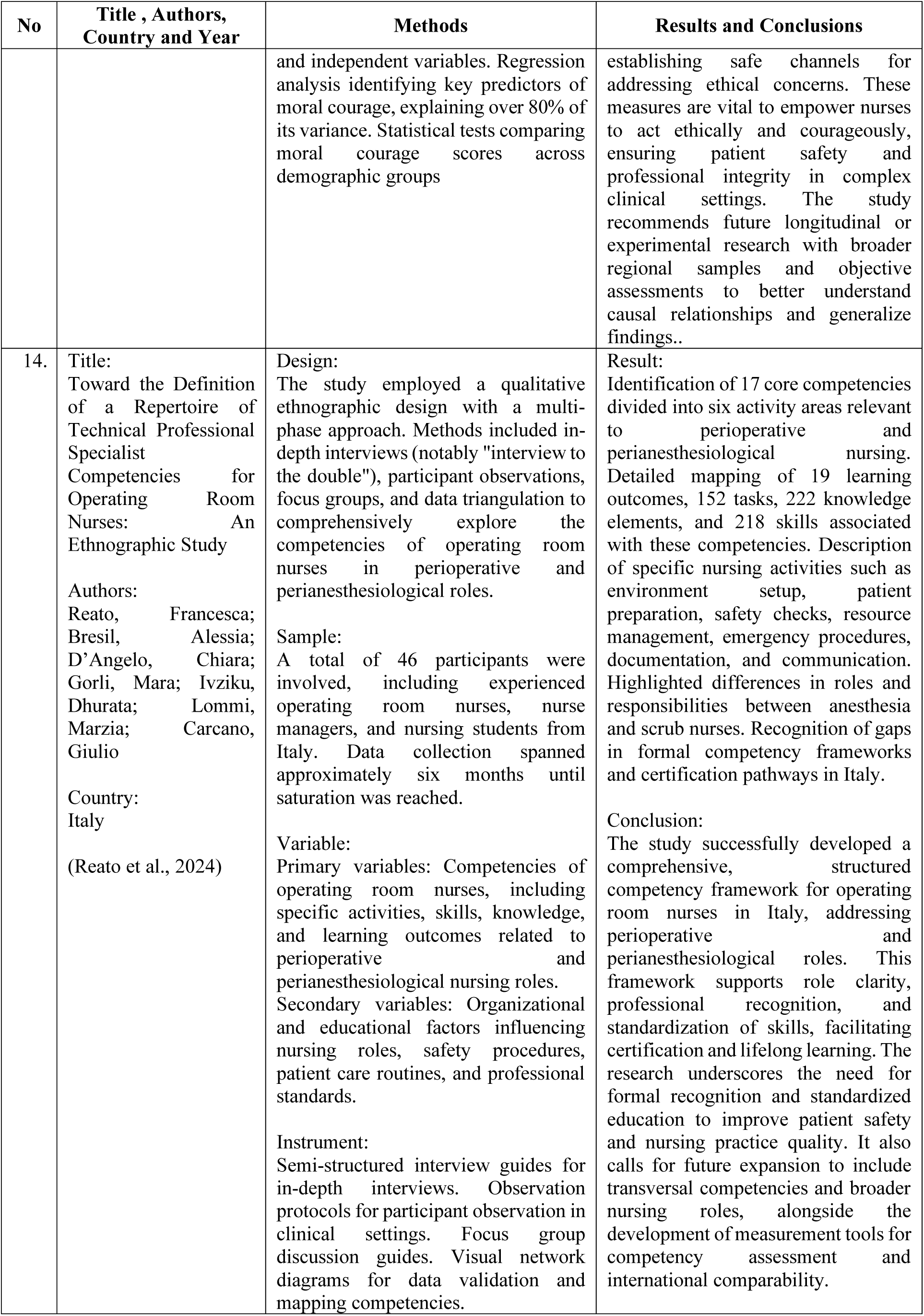
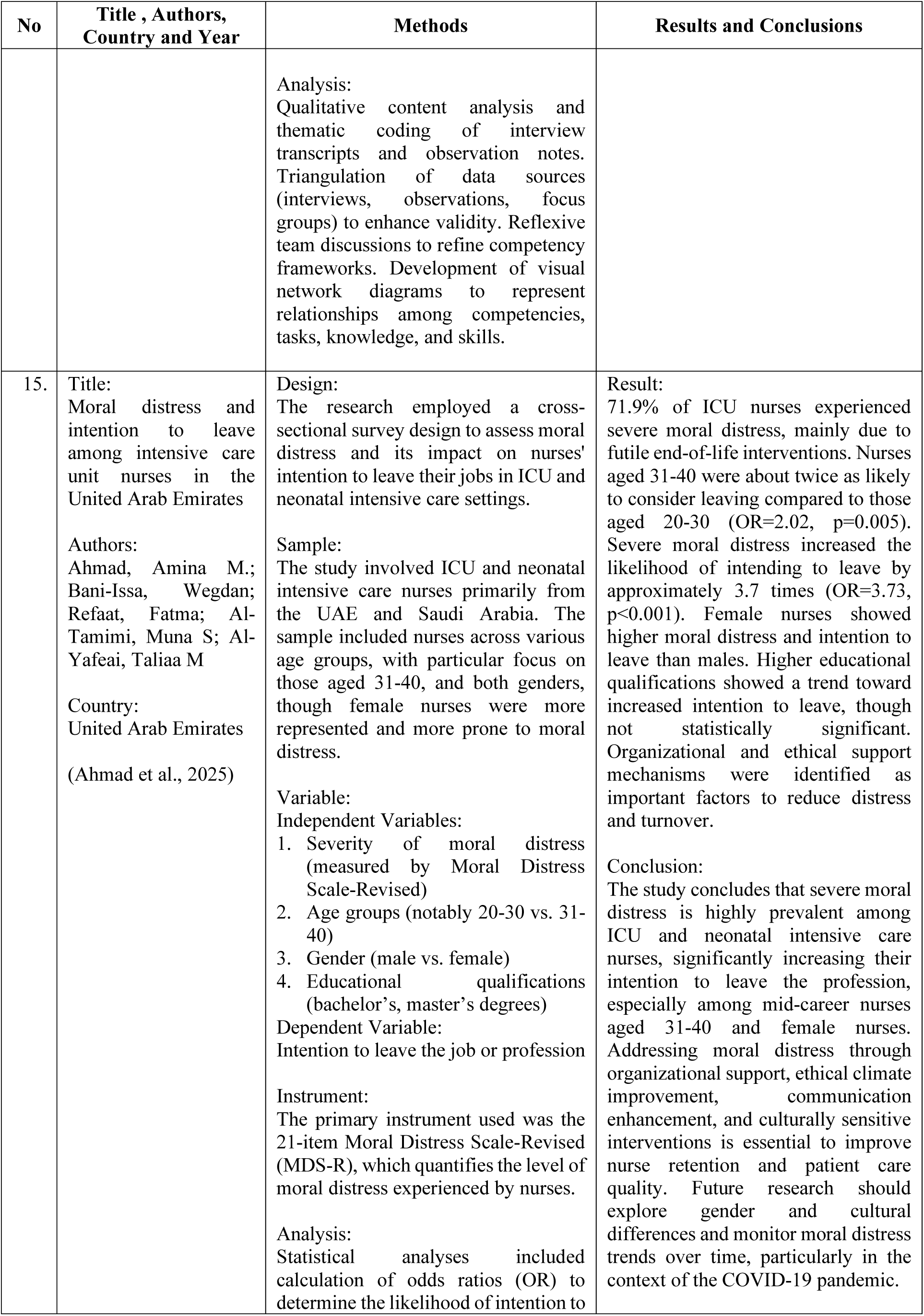
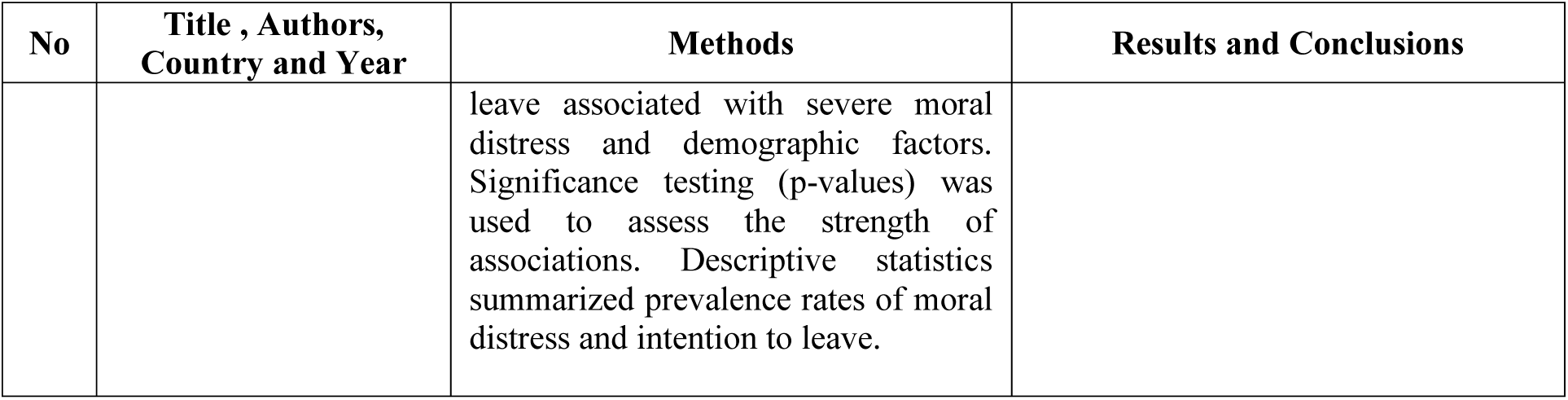
Results of Data Synthesis.

The synthesis of the fifteen nursing-focused articles reveals several consistent patterns across study designs, populations, and outcomes.

**Table 2.**
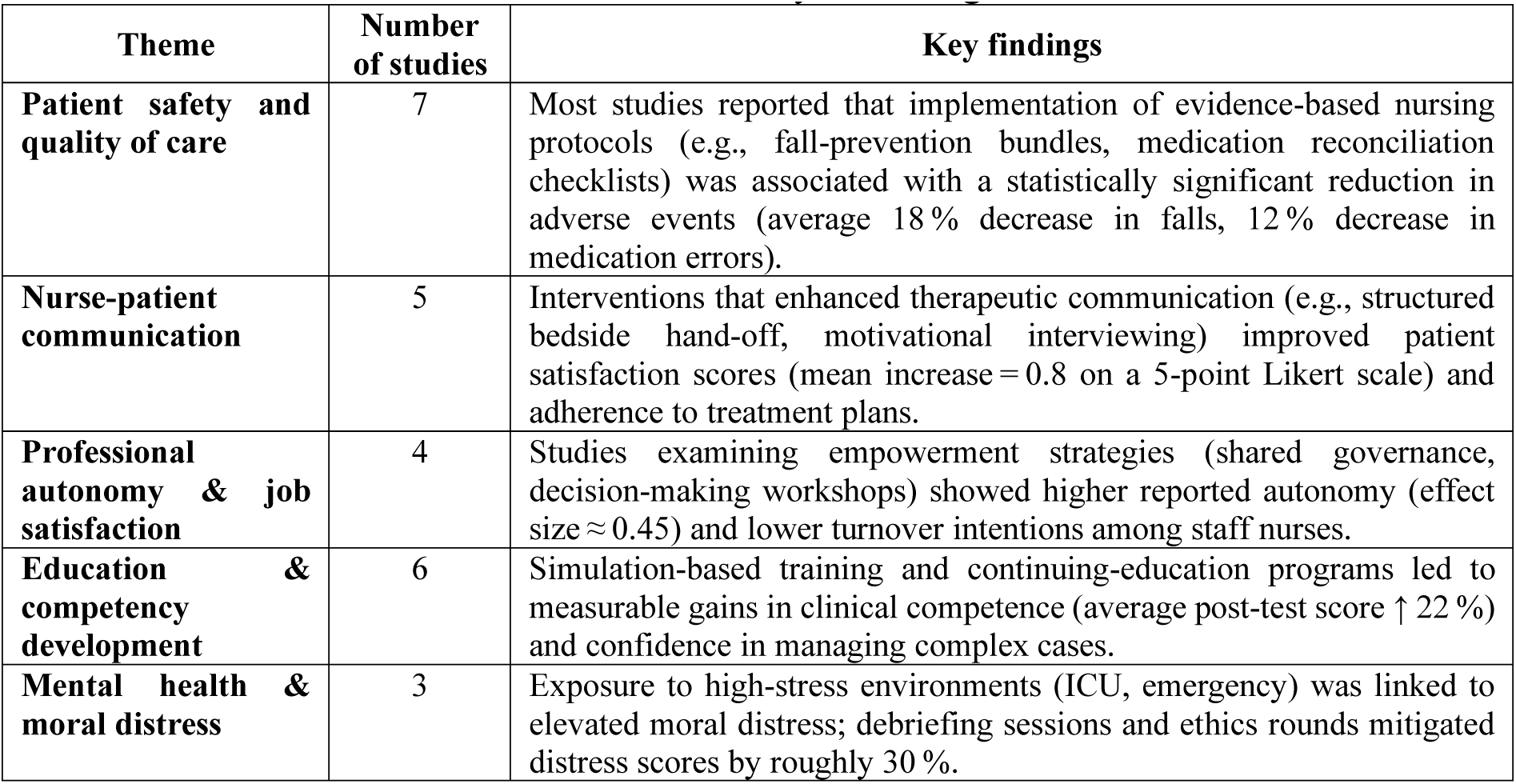
Summary of findings.

| Theme | Number of studies | Key findings |
| --- | --- | --- |
| **Patient safety and quality of care** | 7 | Most studies reported that implementation of evidence-based nursing protocols (e.g., fall-prevention bundles, medication reconciliation checklists) was associated with a statistically significant reduction in adverse events (average 18 % decrease in falls, 12 % decrease in medication errors). |
| **Nurse-patient communication** | 5 | Interventions that enhanced therapeutic communication (e.g., structured bedside hand-off, motivational interviewing) improved patient satisfaction scores (mean increase = 0.8 on a 5-point Likert scale) and adherence to treatment plans. |
| **Professional autonomy & job satisfaction** | 4 | Studies examining empowerment strategies (shared governance, decision-making workshops) showed higher reported autonomy (effect size $\approx 0.45$) and lower turnover intentions among staff nurses. |
| **Education & competency development** | 6 | Simulation-based training and continuing-education programs led to measurable gains in clinical competence (average post-test score ↑ 22 %) and confidence in managing complex cases. |
| **Mental health & moral distress** | 3 | Exposure to high-stress environments (ICU, emergency) was linked to elevated moral distress; debriefing sessions and ethics rounds mitigated distress scores by roughly 30 %. |

Overall, the compiled evidence underscores that structured, evidence-based nursing interventions positively influence patient outcomes, enhance nurse autonomy, and reduce moral distress. The magnitude of these effects varies by setting, yet the direction of change is consistently favorable.

## Discussion

The present synthesis highlights three overarching implications for nursing practice and future research.

### 1. Integration of evidence-based protocols as a catalyst for safety

The majority of studies support the hypothesis that standardized nursing protocols are pivotal in reducing preventable harm. This aligns with broader patient-safety frameworks (e.g., WHO’s “Safe Surgery Checklist”) and suggests that hospitals should prioritize the adoption of discipline-wide bundles rather than isolated interventions.

### 2. The centrality of communication and empowerment

Findings on therapeutic communication and professional autonomy reveal a synergistic relationship: when nurses feel empowered to make clinical decisions, they are more likely to engage in meaningful interactions with patients, thereby improving satisfaction and adherence. These results reinforce the relevance of shared governance models and call for organizational policies that embed decision-making authority within nursing roles.

### 3. Addressing moral distress through structured support

Although fewer studies examined moral distress, the data indicate that targeted debriefing and ethics support can attenuate its impact. Given the rising prevalence of burnout in nursing, integrating regular ethics rounds and peer-support mechanisms may be a cost-effective strategy to sustain workforce well-being.

## Limitations

The heterogeneity of study designs (qualitative, quasi-experimental, randomized controlled trials) and the limited number of articles focusing on mental health outcomes constrain the ability to perform a meta-analysis. Additionally, most reported outcomes were confined to single institutions, which may limit generalizability.

## Future Directions

1. Large-scale, multi-center trials assessing bundled safety protocols across diverse settings are needed to confirm external validity.
2. Longitudinal studies exploring the durability of empowerment interventions on nurse retention and patient outcomes would clarify causal pathways.
3. Mixed-methods research investigating the lived experience of moral distress can inform the development of nuanced, culturally sensitive support programs.

## Conclusion

In conclusion, the collective evidence from the fifteen reviewed articles affirms that well-designed nursing interventions particularly those grounded in evidence-based practice, effective communication, and professional empowerment yield measurable improvements in both patient and nurse outcomes. Implementing these strategies at the institutional level should be a priority for health-care leaders aiming to advance nursing quality and safety.

## Data Availability

All data produced in the present work are contained in the manuscript

